# Splice site and *de novo* mutations can cause mixed dominant negative/gain of function *PLCG2*-associated immune dysregulation with cold urticaria (CU-PLAID)

**DOI:** 10.1101/2024.03.16.24304180

**Authors:** Sophia R. Chou, Alexis C. Bailey, Kathleen Baysac, Andrew J. Oler, Joshua D. Milner, Michael J. Ombrello

## Abstract

**Background:** Phospholipase Cγ2 (PLCγ2) is an important signaling molecule that receives and transmits signals from various cell surface receptors in most hematopoietic lineages. Variants of *PLCG2* cause PLCγ2-associated immune dysregulation (PLAID), a family of conditions that are classified by mutational effect. PLAID with cold urticaria (CU-PLAID) is caused by in-frame deletions of *PLCG2* that are dominant negative at physiologic temperatures but become spontaneously active at sub-physiologic temperatures.

**Objective:** To identify genetic lesions that cause PLAID by combining RNA sequencing of full-length *PLCG2* with whole genome sequencing.

**Methods:** We studied nine probands with antibody deficiency and a positive evaporative cooling test, together with two known CU-PLAID patients and three healthy subjects. Illumina sequencing was performed on full-length *PLCG2* cDNA synthesized from peripheral blood mononuclear cell RNA and whole genome sequencing was used to identify genetic lesions. Novel alternate transcripts were overexpressed in the *Plcg2-*deficient DT40 cell overexpression system. ERK phosphorylation was quantified by flow cytometry with and without BCR crosslinking.

**Results:** Two probands expressed novel alternative transcripts of *PLCG2* with in-frame deletions. The first, expressing *PLCG2* without exons 18-19, carried a splice site mutation in intron 19. The second, expressing *PLCG2* without exons 19-22, carried a 14kb *de novo* deletion of *PLCG2*. DT40 cells overexpressing the exon 18-19 or exon 19-22 deletions failed to phosphorylate ERK in response to BCR crosslinking.

**Conclusion:** In addition to autosomal dominant genomic deletions, *de novo* deletions and splice site mutations of *PLCG2* can also cause CU-PLAID. All of these can be identified by cDNA-based sequencing.

**Capsule Summary:** By identifying both the first *de novo* and splice site variants to cause *PLCG2*-associated immune dysregulation with cold urticaria (CU-PLAID), we demonstrate the diagnostic utility of *PLCG2*-specific RNA-sequencing.

## Introduction

PLCγ2 is an important signaling molecule in a range of leukocytes, with clear importance in B lymphocytes, NK cells, mast cells and neutrophils [1]. Variation in *PLCG2* causes PLCγ2-associated immune dysregulation (PLAID), a family of conditions that can be subdivided by mutational effect [2]. We first observed in-frame deletions that lead to a mixed dominant negative (DN) loss of function (LOF) with cold-induced gain of function (GOF) were observed. These deletions caused autosomal dominant cold urticaria with antibody deficiency and increased susceptibility to autoimmunity, atopy, and infection in three unrelated families; this was originally called PLAID [3]. Next, a series of GOF variants were described in patients and families with humoral abnormalities and a range of inflammatory features, originally called autoinflammatory PLAID (APLAID) [2,4]. Most recently, two studies described LOF mutations of *PLCG2* in individuals with immune dysregulation, including a subset with autosomal dominant recurrent herpesvirus infections and NK cell immunodeficiency, with or without humoral defects [2,5]. We observed that the features of immune dysregulation in these three mutational classes largely overlapped [2], except for cold urticaria induced by evaporative cooling, which has to date been universally and exclusively observed in the patients with in-frame deletions. For clarity, we propose that this condition be referred to as PLAID with cold urticaria (CU-PLAID).

Each of the three published families with CU-PLAID bears a unique in-frame genomic deletion that excludes either exon 19 (Δ19) or exons 20-22 (Δ20-22) from the autoinhibitory domain of PLCγ2 [3]. These mutations produce a paradoxical combination of hypomorphic cellular activity at physiologic temperatures and constitutive activation at cool temperatures, leading to the distinctive clinical combination of humoral immune deficiency and cold urticaria induced by evaporative cooling. The deletions that cause PLAID were identified by manual screening of *PLCG2* transcripts with a cumbersome and time-consuming combination of PCR, Sanger sequencing, and transcriptomic skip assays that frequently failed to identify genomic deletions [3,6]. This approach is highly dependent on primer placement and failed to identify causative mutations in a group of phenotypically identical patients [6]. To streamline the screening process while addressing the low abundance of *PLCG2* transcripts, we employed gene-specific RNA-sequencing (RNA-seq) of *PLCG2* and whole genome sequencing (WGS) to search for causative genetic lesions of *PLCG2* in subjects with clinical features consistent with CU-PLAID.

## Results/Discussion

Our study included nine probands with lifelong cold urticaria associated with a positive evaporative cooling test and antibody deficiency. The study included one member of a multigenerational family with autosomal dominant cold urticaria (Proband 1) and eight sporadic cases, one of which was a child who was symptomatic since birth but whose parents were known to be asymptomatic (Proband 2; Figure 1). Three healthy controls and two known PLAID patients were used as negative and positive controls respectively. RNA was obtained by isolating peripheral blood mononuclear cells from patients by gradient centrifugation and RNeasy kits (Qiagen, Germantown, MD) were used to isolate RNA. Full-length *PLCG2* cDNA was synthesized by reverse transcription with SuperScript III Reverse Transcriptase (Thermo Fisher Scientific, Waltham, MA) and a *PLCG2*-specific primer that recognizes a region of the 3’ untranslated region that is conserved in almost all *PLCG2* transcripts (Supplementary Table 1). Full-length *PLCG2* was amplified from cDNA by long-range PCR with TaKaRa LA Taq (Takara Bio, San Jose, CA) and custom primers (Supplementary Table 1). Sequencing libraries were generated using Illumina Nextera kits and were sequenced with 150bp paired end reads on a MiSeq instrument (Illumina, San Diego, CA). Reads were aligned to the *PLCG2* cDNA sequence (hg19) using STAR and parsed for alternate transcripts containing deletions.

**Figure 1.**
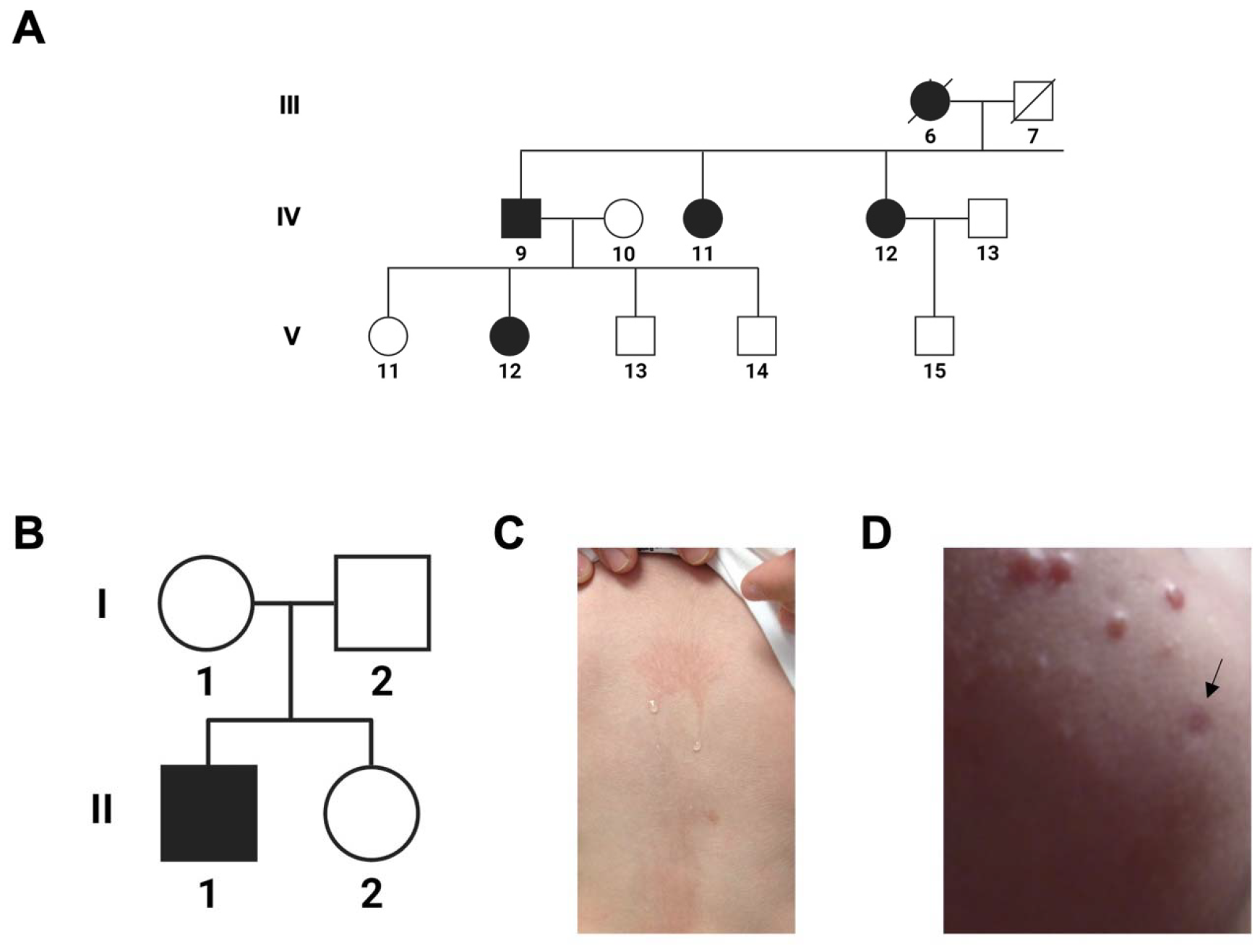
Patient Pedigree and Clinical Manifestations. Proband 1 **(A**, subject V-12) is a member of a family displaying multigenerational, autosomal dominant cold induced urticaria and immune dysregulation. A full pedigree of this family and their clinical features have been described by Gandhi et al.^7^. Proband 2 (**B**, subject II-1) was sporadically affected by cold urticaria since birth that could be induced by evaporative cooling **(C)**. He developed scattered granulomata **(D)**, one of which was biopsied (black arrow) and demonstrated well-formed granulomata with giant cells and numerous histocytes highlighted by CD68. Proband 2 was previously described in a report by Aderibigde et al.^6^.

Through *PLCG2*-specific RNA-seq, we observed four probands who had one or more exon-skipping transcripts that accounted for > 10% of reads (Figure 2A). Two of these individuals were found to express transcripts with in-frame deletions of the regulatory region of PLCγ2 that were not observed in healthy subjects. Proband 1 had an exon 18-19 deletion (Δ 18-19), an exon 19-22 deletion (Δ19-22) and Δ 19, whereas proband 2 had an exon 19-22 deletion (Δ19-22; Figure 2B). When the exon-skipping cDNA sequences of these two subjects were aligned to *PLCG2* and compared, the boundaries of Δ19 and Δ19-22 were well-defined. The Δ18-19, Δ19 and Δ19-22 junctions were each confirmed by conventional sequencing (Supplementary Figure 1).

**Figure 2.**
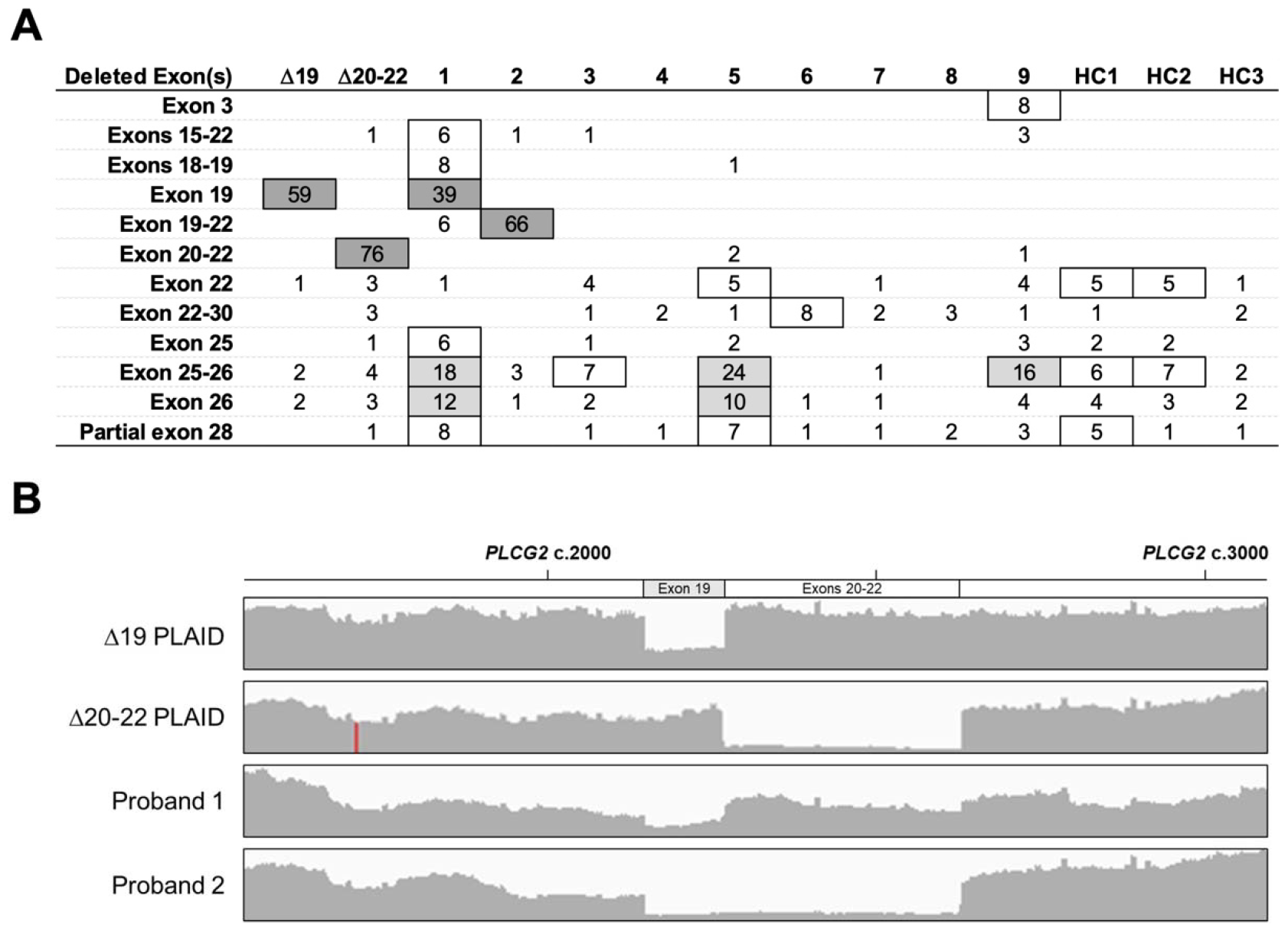
RNA sequencing of full length *PLCG2* identifies landscape of alternative transcripts. *PLCG2*-specific RNA sequencing reads were aligned to the canonical transcript (NM_002661.5) using STAR RNA-seq aligner. Exon-skipping alternative transcripts composing ≥ 5% of total reads in at least 1 individual are displayed in **Panel A**. The sequencing reads of subjects with high proportion (> 30%, dark grey) alternative transcripts were aligned and visualized with Integrative Genomics Viewer **(B)**.

We performed trio WGS on these two subjects and their parents. In the first patient, who had a combination of Δ18-19, Δ19 and Δ19-22 transcripts, we discovered a splice site mutation located in intron 19 (*PLCG2* c.2298+5N>T). In the second with the Δ19-22 transcript, WGS revealed a 14kb *de novo* deletion of *PLCG2* that spanned from intron 18 to intron 22, causing the deletion of exons 19-22 (chr16.81944583_81958780del; Figure 3A). The *de novo* mutation was of particular interest, as it provided the first opportunity to directly investigate the origin of a CU-PLAID causing *PLCG2* deletion. Two of the previously published CU-PLAID deletions were observed to have breakpoints that intersected with Alu repetitive elements on both ends. [7] (Supplementary Figure 2). The alignment of these elements strongly suggests that they resulted from an Alu-mediated deletional event. In the case of the patient with the *de novo* mutation, there were no repetitive elements intersecting with the deletional breakpoints (Figure 3B). An alternative explanation may lie in the fact that *PLCG2* is situated in a region of chromosome 16 that is a documented hotspot for the development of double strand DNA breaks and *de novo* mutations [8]. Moreover, the deletional rate of this hotspot is proportional to maternal age, with the highest rates observed in women over the age of 36 years [8]. Proband 2’s deletion arose on the maternally transmitted chromosome and his mother was within this age range, which together reinforce the plausibility this mechanism. This suggests that screening of patients with antibody deficiency and cold urticaria, even when not familial, may be useful in the diagnosis of CU-PLAID.

**Figure 3.**
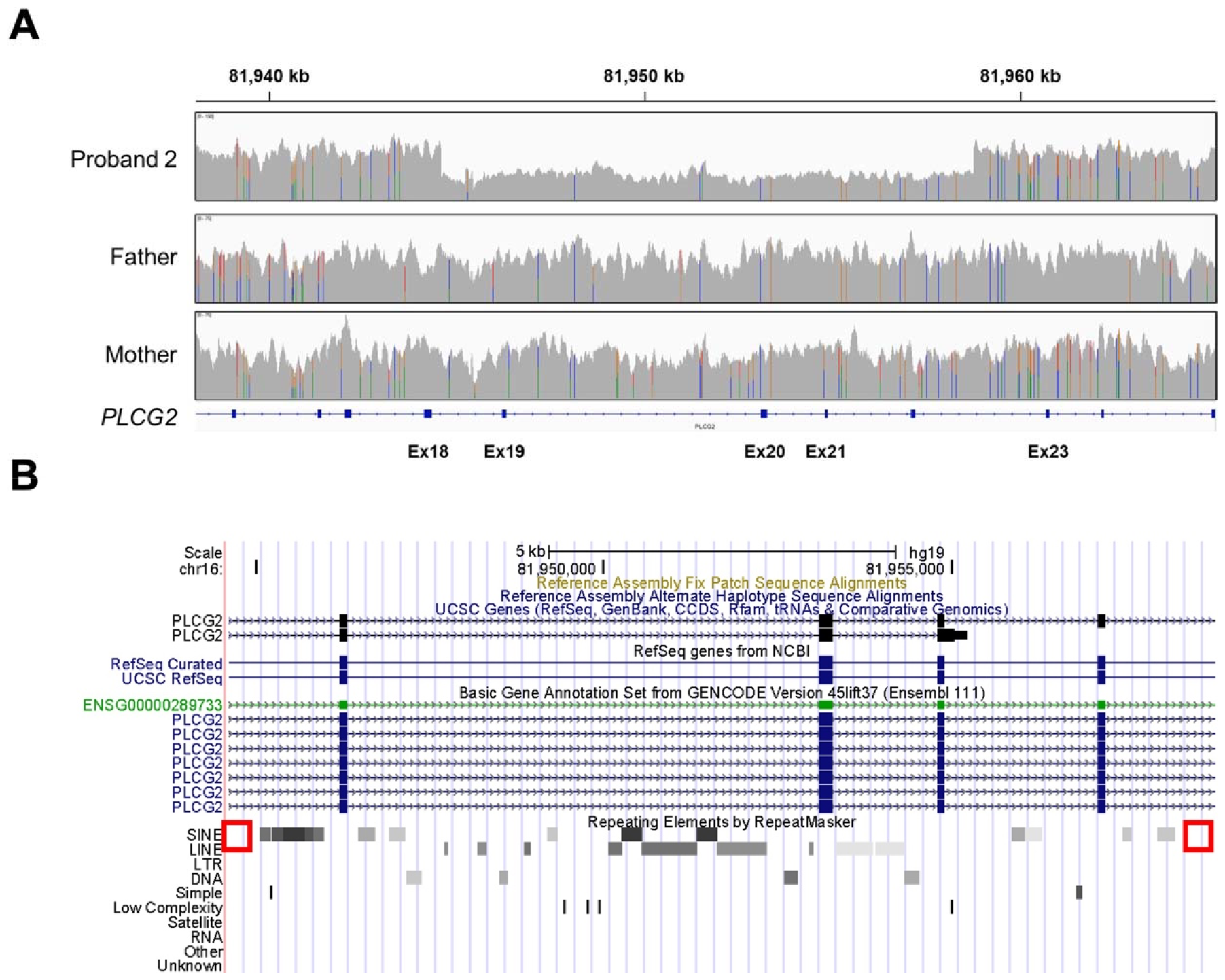
*De novo* deletion of *PLCG2* causes novel CU-PLAID causing alternative transcript. Whole genome sequencing reads from proband 2 and his parents illustrate a deletion in the proband that is not observed in either parent **(A)**. In **panel B**, the deleted segment from proband 2 is displayed on the University of Santa Cruz Genome Browser, revealing an absence of intersecting Alu or other short interspersed nuclear elements at the deletional breakpoints (red boxes).

These mutations reflect the first splice site mutation and *de novo* mutations to cause CU-PLAID, respectively. To evaluate the functional effect of the new exon-skipping transcripts, we used the *Plcg2*^*-/-*^ DT40 cell overexpression system, as previously described [2]. In doing so, we observed that cells transfected with the Δ18-19 or Δ19-22 forms of *PLCG2* failed to phosphorylate ERK in response to BCR crosslinking (Figure 4), consistent with cells overexpressing the published Δ19 and Δ20-22 forms of *PLCG2*. This study identified two novel *PLCG2* mutations that cause CU-PLAID, both of which encode a new alternate transcript, expanding our knowledge of the disease.

**Figure 4.**
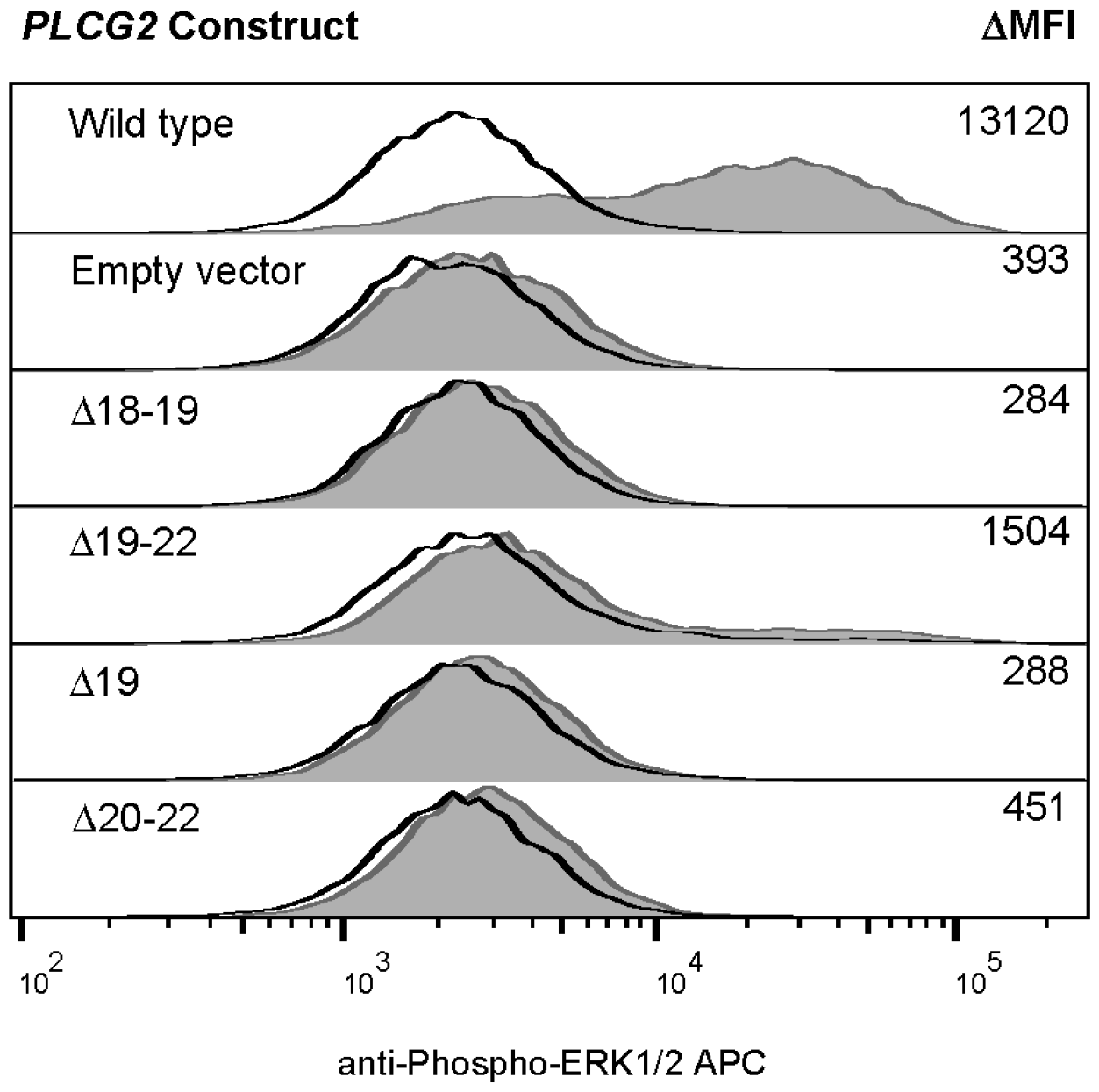
Novel alternate transcripts of *PLCG2* lead to failure of ERK phosphorylation. Using the *Plcg2*^*-/-*^ DT40 overexpression system, the effects of published (Δ19, Δ 20-22)^3^ and novel (Δ 18-19, Δ 19-22) in frame deletions of *PLCG2* on unstimulated (black tracing) and IgM-stimulated (grey fill) ERK phosphorylation were assayed by flow cytometry, as previously described^2^. Δ 19, exon 19 skipped *PLCG2;* Δ 20-22, exon 20-22 skipped *PLCG2;* Δ 18-19, exon 18-19 skipped *PLCG2;* Δ 19-22, exon 19-22 skipped *PLCG2*.

*PLCG2* transcripts are relatively low abundance in PBMC, which complicates identification of mutations among transcript-wide cDNA libraries. In addition, detection of exon-skipping *PLCG2* transcripts by direct Sanger sequencing is obscured by the presence of multiple different species of the transcript, including the canonical transcript produced by the non-causative (wild type) allele and the group of alternative transcripts that are produced by either allele. This can be further compounded with splice-site mutations, which may produce a plurality of alternatively spliced transcripts, making it nearly impossible to identify the mutation using PCR or Sanger sequencing. Therefore, we chose to use gene-specific reverse transcription of PBMCs to generate full-length *PLCG2* cDNA, coupled to next generation sequencing. This unbiased strategy allowed us to characterize the full range of *PLCG2* transcrip*t* variation from primary leukocytes.

Our novel cDNA-based method of detecting alternate transcripts of *PLCG2* allowed us to establish that 1) a single splice-site substitution can produce the same autosomal dominant phenotype as a genomic deletion, 2) *de novo* mutations can lead to non-familial cases of CU-PLAID, and 3) cDNA-based sequencing can substantially improve diagnostic accuracy of CU-PLAID. This approach is applicable to other genetic diseases of the immune system that are caused by alternative transcripts, such as the NEMO Δ exon 5 autoinflammatory syndrome [9].

## Data Availability

All data produced in the present study are available upon reasonable request to the authors

## Acknowledgements

This study was funded by the Intramural Research Program of the National Institute of Arthritis and Musculoskeletal and Skin Diseases (Z01-AR041198). Additional support was provided by the Division of Intramural Research, National Institute of Allergy and Infectious Diseases (Z01-AI001098). This work utilized the resources of the Genomics Technology Section and the Flow Cytometry Section of the Intramural Research Program of NIAMS. The work utilized the computational resources of the NIH high-performance computing cluster Biowulf (http://hpc.nih.gov).

## References

1. Jackson JT, Mulazzani E, Nutt SL, et al. The role of PLCgamma2 in immunological disorders, cancer, and neurodegeneration. J Biol Chem. 2021 Aug;297(2):100905.

2. Baysac K, Sun G, Nakano H, et al. PLCG2-associated immune dysregulation (PLAID) comprises broad and distinct clinical presentations related to functional classes of genetic variants. J Allergy Clin Immunol. 2024 Jan;153(1):230–242.

3. Ombrello MJ, Remmers EF, Sun G, et al. Cold urticaria, immunodeficiency, and autoimmunity related to PLCG2 deletions. N Engl J Med. 2012 Jan 26;366(4):330–8.

4. Zhou Q, Lee GS, Brady J, et al. A hypermorphic missense mutation in PLCG2, encoding phospholipase Cgamma2, causes a dominantly inherited autoinflammatory disease with immunodeficiency. Am J Hum Genet. 2012 Oct 5;91(4):713–20.

5. Alinger JB, Mace EM, Porter JR, et al. Human PLCG2 haploinsufficiency results in a novel natural killer cell immunodeficiency. J Allergy Clin Immunol. 2024 Jan;153(1):216–229.

6. Aderibigbe OM, Priel DL, Lee CC, et al. Distinct Cutaneous Manifestations and Cold-Induced Leukocyte Activation Associated With PLCG2 Mutations. JAMA Dermatol. 2015 Jun;151(6):627–34.

7. Gandhi C, Healy C, Wanderer AA, Hoffman HM. Familial atypical cold urticaria: description of a new hereditary disease. J Allergy Clin Immunol. 2009;124(6):1245–1250. doi:10.1016/j.jaci.2009.09.035

8. Goldmann, J.M., Seplyarskiy, V.B., Wong, W.S.W. et al. Germline de novo mutation clusters arise during oocyte aging in genomic regions with high double-strand-break incidence. Nat Genet 50, 487–492 (2018). 10.1038/s41588-018-0071-6

9. Lee Y, Wessel AW, Xu J, et al. Genetically programmed alternative splicing of NEMO mediates an autoinflammatory disease phenotype. J Clin Invest. 2022 Mar 15;132(6).

