## Supplementary Appendix for "Splice site and *de novo* mutations can cause mixed dominant negative/gain of function *PLCG2*-associated immune dysregulation with cold urticaria (CU-PLAID)"

**Supplementary Table 1. Primer sequences for amplifying full length *PLCG2* cDNA**

| **Primer function** | **Forward** | **Reverse** |
| --- | --- | --- |
| **Reverse transcription of full length *PLCG2* cDNA** |  | GCACGTTCTCCTACATGCAA |
| **PCR amplification of full length *PLCG2* cDNA** | GCCAGCTTCCTGATTTCTCC | GCACGTTCTCCTACATGCAA |

**Supplementary Figure 1. Validation of alternative *PLGC2* transcript variants by conventional sequencing.** The breakpoints of exon-skipping alternative transcripts of *PLCG2* were examined by conventional sequencing using previously published primer pairs [1].

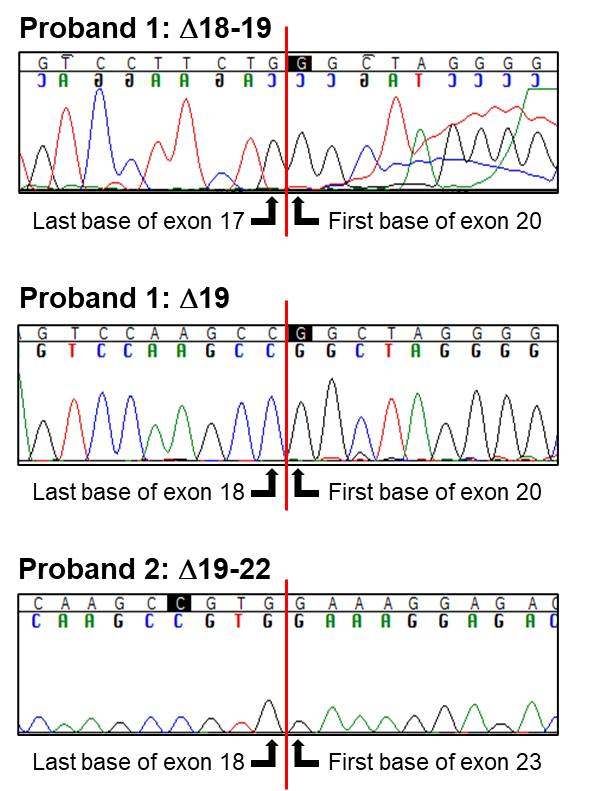

**Supplementary Figure 2. Microhomology of Alu elements at breakpoints of published Δ19 *PLCG2* alleles**. The deleted segments from Family 1 **(A)** and Family 3 **(B)** from Ombrello MJ et al. [1] were visualized on the University of Santa Cruz Genome Browser, revealing the presence of intersecting short interspersed nuclear elements (Alu elements) at each deletional breakpoint (red boxes). In **Panels C and D**, the family-specific deleted allele sequences are aligned with the reference sequences of *PLCG2* intron 18 and intron 19, revealing microhomology of Alu elements. Pink indicates regions of complete homology of Alu elements between strands surrounding the breakpoints. Blue indicates identity between the familial sequence and the reference sequence.
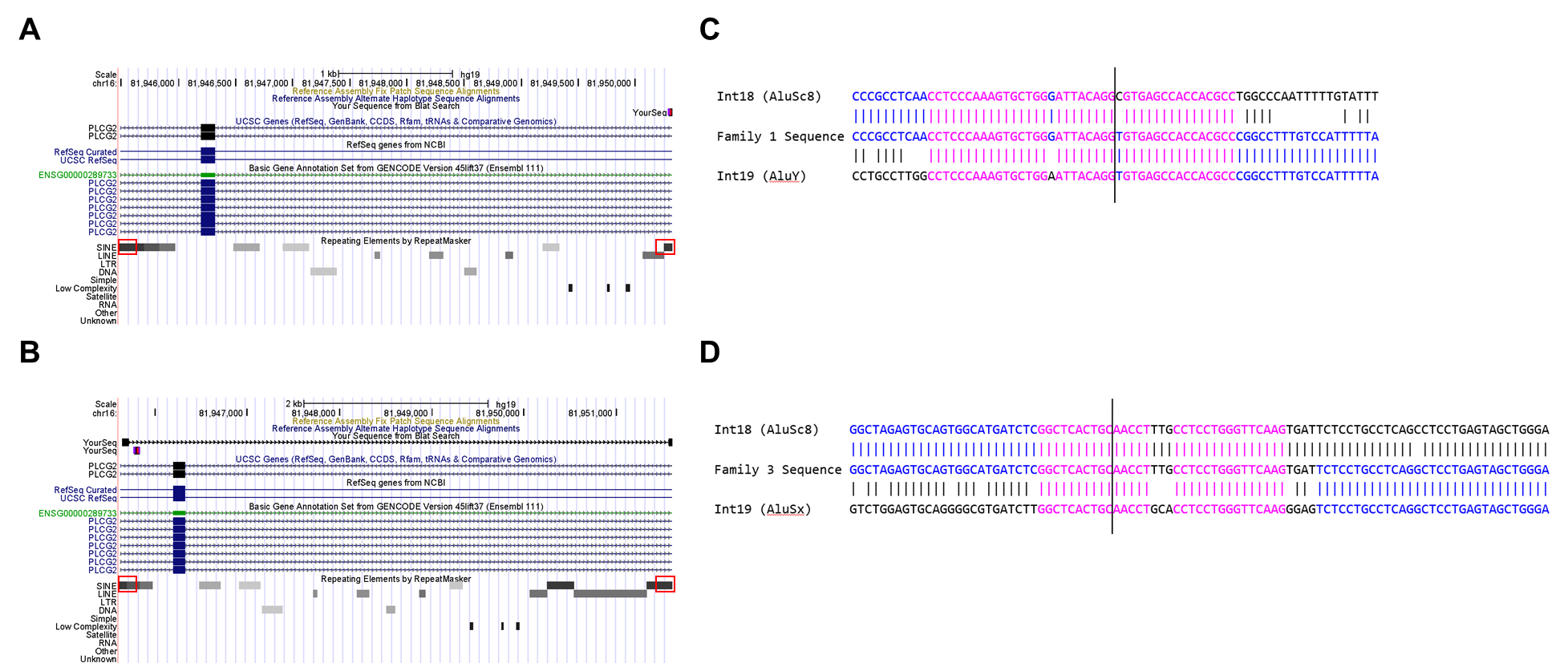

**Supplementary References**

1. Ombrello MJ, Remmers EF, Sun G, et al. Cold urticaria, immunodeficiency, and autoimmunity related to *PLCG2* deletions. *N Engl J Med*. 2012 Jan 26;366(4):330-8.
